# COVIDStrategyCalculator: A software to assess testing- and quarantine strategies for incoming travelers, contact person management and de-isolation

**DOI:** 10.1101/2020.11.18.20233825

**Authors:** Wiep van der Toorn, Djin-Ye Oh, Max von Kleist, on behalf of the working group on SARS-CoV-2 Diagnostics at RKI

## Abstract

While large-scale vaccination campaigns against SARS-CoV-2 are rolled out at the time of writing, non-pharmaceutical interventions (NPIs), including the isolation of infected individuals, and quarantine of exposed individuals remain central measures to contain the spread of SARS-CoV-2. Strategies that combine NPIs with innovative SARS-CoV-2 testing strategies may increase containment efficacy and help to shorten quarantine durations.

We developed a user-friendly software-tool that implements a recently published stochastic within-host viral dynamics model that captures temporal attributes of the viral infection, such as test sensitivity, infectiousness and the occurrence of symptoms. Based on this model, the software allows to evaluate the efficacy of user-defined, arbitrary NPI and testing strategies in reducing the transmission potential in different contexts. The software thus enables decision makers to explore NPI strategies and perform hypothesis testing, e.g. with regards to the utilization of novel diagnostics or with regards to containing novel virus variants.

## Introduction

Non-pharmaceutical interventions (NPIs), are important tools to prevent SARS-CoV-2 transmission and contain the spread of novel variants. NPIs consist of quarantine, isolation and diagnostic testing of (potentially) infected individuals. The term ‘quarantine’ refers to the separation of people who are at risk of being infected with SARS-CoV-2 due to potential exposure, but whose infection status is unknown. Examples include the management of incoming travelers from high-risk areas, or the management of individuals who have been in contact with confirmed cases. The term ‘isolation’, refers to the separation of individuals with a confirmed infection. NPI strategies may also combine quarantine, isolation and SARS-CoV-2 testing to improve efficacy, or shorten quarantine durations.

For quarantine, WHO recommends a length of 14 days^1^ and for isolation, a length of at least 13 days^2^. However, at the national, and sub-national or institutional level different strategies are often implemented. This may be due to perceived socioeconomic pressure^3^ or staffing concerns in the healthcare systems^4^. In these settings, testing is frequently used to shorten the duration of quarantine and/or isolation. Given that antigen-based rapid diagnostic tests (RDT) are being used increasingly^5^, strategies that are based on combined testing and quarantine/isolation criteria may gain even more momentum in the near future.

To enable the design and evaluate the efficacy of NPI strategies in preventing the risk of onwards transmission, we present the COVIDStrategyCalculator software. The software implements a stochastic intra-host SARS-COV-2 dynamics model, presented in an associated article^6^, to assess arbitrary, user-defined, NPI strategies ‘on the fly’. The software focuses on three common scenarios in policy making: (i) contact person management, (ii) quarantine of incoming travelers and (iii) isolation strategies.

The COVIDStrategyCalculator software can be run in the browser (https://COVIDStrategyCalculator.github.io), or be downloaded as an offline version for Mac, Linux and Windows (https://github.com/COVIDStrategyCalculator/COVIDStrategyCalculator). The software allows full flexibility with regards to parameter choices of the underlying model, that, for example, determine the time-course of infection (e.g. the average “incubation time” or the “time of infectiousness”), the proportion of asymptomatic cases, the test sensitivity and much more. However, a set of default parameters has been derived in an associated article^6^ that synthesizes the current knowledge on within-host infection dynamics of SARS-CoV-2 and temporal test sensitivity.

NPI modelling studies^7-12^ often present particular precomputed NPI scenarios. This leaves policy makers the choice to either accept all assumptions of a pre-computed scenario, implementing it into a national guideline, or to try to interpolate between the precomputation-assumptions and the actual situation faced. Both approaches leave a knowledge gap that precludes the analysis of the exact NPI strategy in question. The COVIDStrategyCalculator fills this void by allowing the user to define an arbitrary NPI strategy and to evaluate its efficacy in preventing onward transmission. The COVIDStrategyCalculator therefore facilitates the rational, evidence-based design of non-pharmaceutical control strategies.

## Results

### Intra-host viral dynamics

The COVIDStrategyCalculator analytically solves a stochastic transit compartment model (Fig. 1A) of the SARS-CoV-2 infection time course. The underlying transit compartment model consists of different phases that resemble relevant attributes of the infection: I.e. whether the virus is (i) detectable, the individual (ii) has symptoms and (iii) may be infectious. In an associated article^6^, we describe the mathematical details of the model, and exemplify the estimation of default parameters that capture clinically observed temporal changes and variability in test sensitivities, incubation- and infectious periods, as well as times to symptom onset.

**Figure 1.**
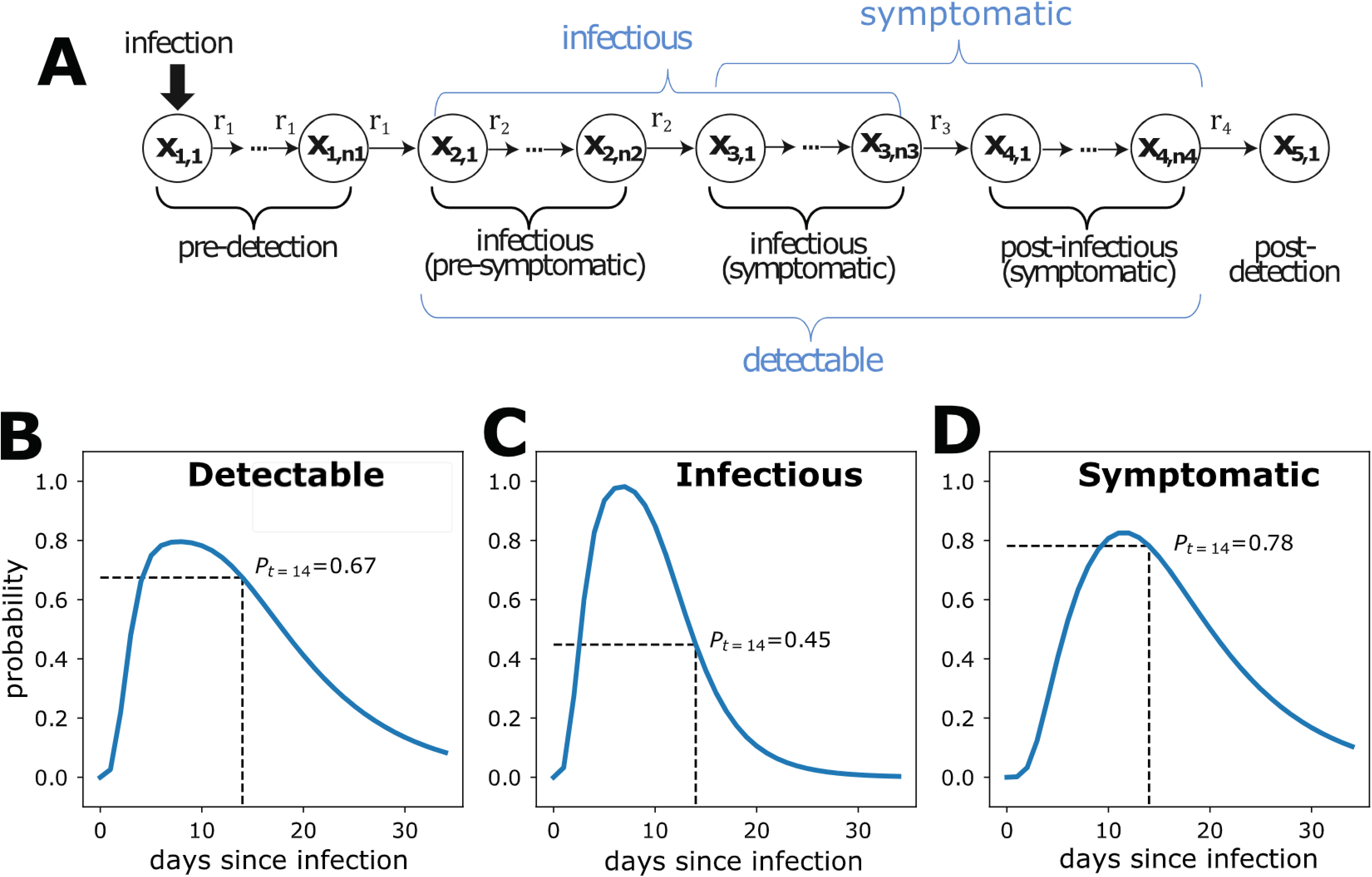
Intra-host viral dynamics. **A.** Stochastic transit compartment model of intra-host viral dynamics. Model details are provided in an associated article^6^. **B-D**. Model simulation with regards to time-dependent **B** virus detectability by PCR, **C** infectiousness and **D** the probability of experiencing symptoms.

In essence, for user-defined parameters, non-pharmaceutical interventions (‘symptom screening’ and testing) and initial states (exemplified below in paragraph *Configuring and simulating an NPI strategy*) COVIDStrategyCalculator internally computes the probability over time of the aforementioned attributes (i-iii) based on the stochastic intra-host model (Fig. 1A). An example is given in Figure. 1B-D: In this simulation, infection occurred at time *t*_O_ = 0. Figure 1B shows the probability that the virus is detectable by PCR (= temporal test sensitivity); Fig. 1C depicts the probability that the individual is infectious (= able to disseminate infectious particles) and Fig. 1D depicts the probability that the individual is symptomatic. For any time point, e.g. 14 days post-infection (black dashed line in Fig. 1B-D), it is possible to compute the virus detection probability, the probability that an individual is infectious and has symptoms. The detection probability (Fig. 1B) and the probability to have symptoms (Fig. 1C) are relevant for different non-pharmaceutical interventions (NPI), e.g. diagnostic testing and ‘symptom screening’, whereas the probability to be infectious (Fig. 1C) determines whether individuals may transmit the virus, and is thus used to compute the efficacy of NPIs in reducing the transmission potential.

### Software outputs

The COVIDStrategyCalculator allows to compute the efficacy of user-defined NPI strategies. Efficacy refers to reducing the transmission potential emanating from an individual (= ‘risk reduction’).

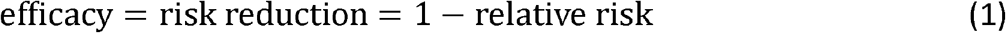

Mathematically, the ‘relative risk’ is computed as the *residual transmission risk* that remains after a user-defined NPI strategy, relative to the transmission risk in a baseline scenario, where no NPIs are imposed (an example is given in the associated article^6^, Fig. 2 therein).

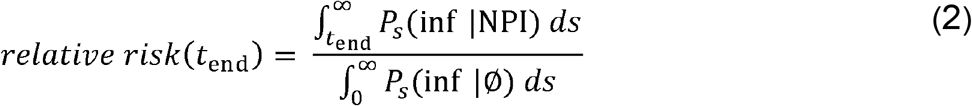

where the nominator integrates over the conditional probability of being infectious after release from an NPI (e.g. quarantine) at time t_end_, whereas the nominator integrates over the probability of being *infectious* in the case where individuals had **not** been isolated, or put into quarantine (baseline risk). Notably, the *relative risk* metric above is independent of the initial prevalence (see also van der Toorn et al.^6^ for details). Importantly, in the equation above we assume no additional (e.g. behavioral) differences between the two settings.

**Figure 2.**
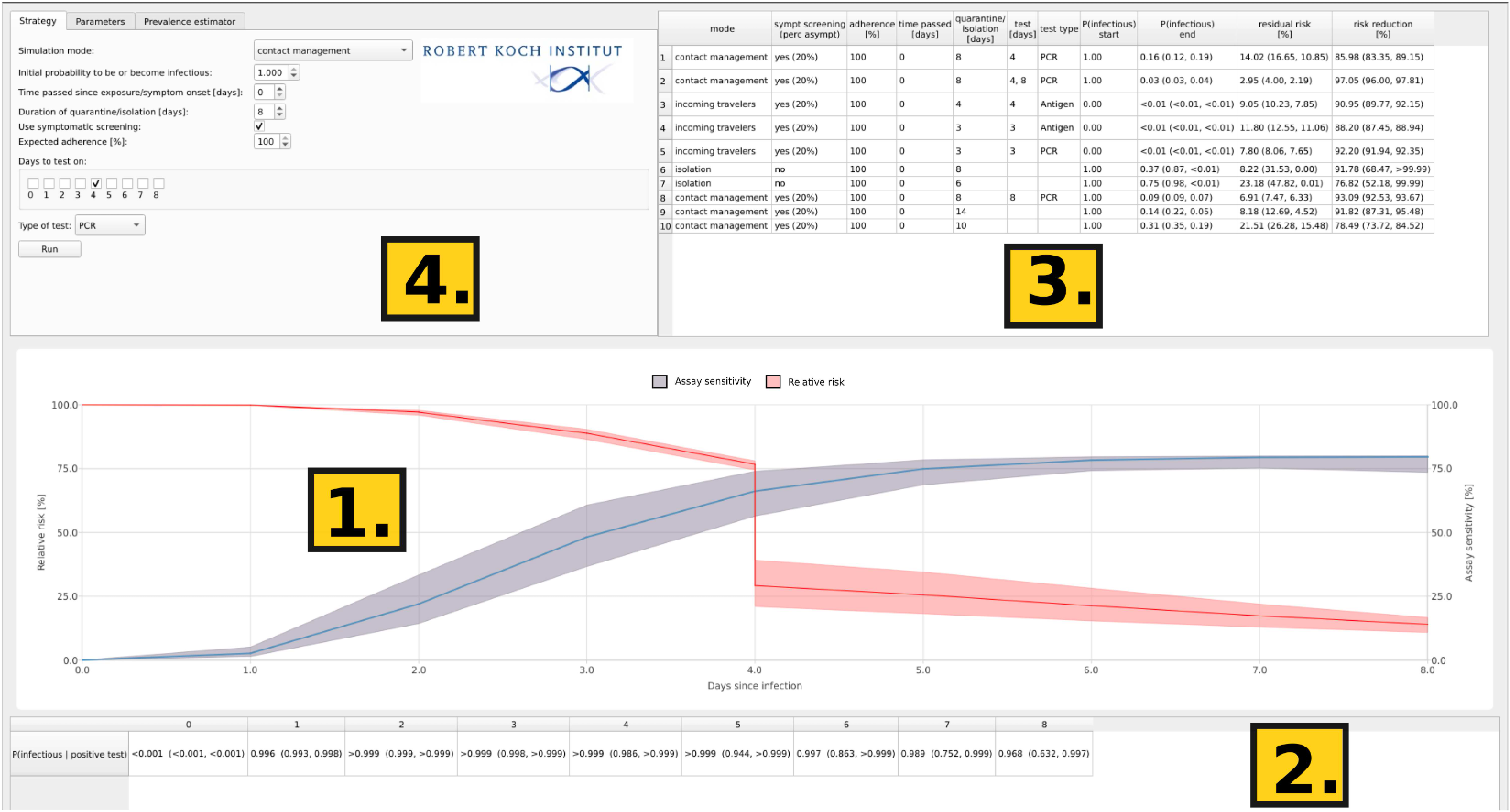
Screenshot of COVIDStrategyCalculator. The main window consists of four components: Time course trajectories (1), test efficacy reports (2), a result log (3) and user input (4.). Reported results include the simulation settings, the pre- & post-procedure probability of infectiousness, as well as the efficacy estimates, depicted as ‘% relative risk’ and the ‘% risk reduction’.

When simulating a user-defined NPI strategy, the profile of the % relative risk will be depicted together with the time-dependent diagnostic assay sensitivity *P*(positive test | infected) as shown in Figure 2 (1.). The latter is intended to visually ease the selection of times to perform diagnostic tests together with the table below the graphic Fig. 2 (2.), which assesses the assays’ ability to filter out *infectious* individuals P(infectious | positive test). We differentiate between the two quantities because PCR assays may allow virus detection for a prolonged time after symptom onset, during which the secreted virus often is not *infectious* anymore. The Table in Fig. 2 (3.) logs the summary of the conducted simulations and outputs the following numeric values; (i) the probability to be infectious *at the time point* where the quarantine or isolation ends 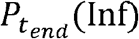, (ii) the ‘% relative transmission risk’ as defined above and (iii) the ‘% risk reduction’ (= efficacy of the NPI). For each metric (Fig. 2 (2. & 3.)), three values are reported which indicate the result in the case of a ‘typical’ infection (typical parameters as outlined below), as well as an uncertainty range based on the provided lower and upper extreme parameter values. For the graphics (Fig. 2 (1.)), the uncertainty range (lower and upper extreme dynamics) is visualized by the shaded area. We will further discuss the parameters of the software and the underlying modeling assumptions by describing how to use the COVIDStrategyCalculator.

In Fig. 2 (4.) the user can configure an arbitrary NPI strategy as outlined below.

### Software utilization

Figure 3A shows a zoom of the opening window: The different tabs allow to evaluate a strategy (1.), set parameters (2.) or perform a prevalence estimation (3). In field (4.), the user can select between the different modes of the software.

**Figure 3.**
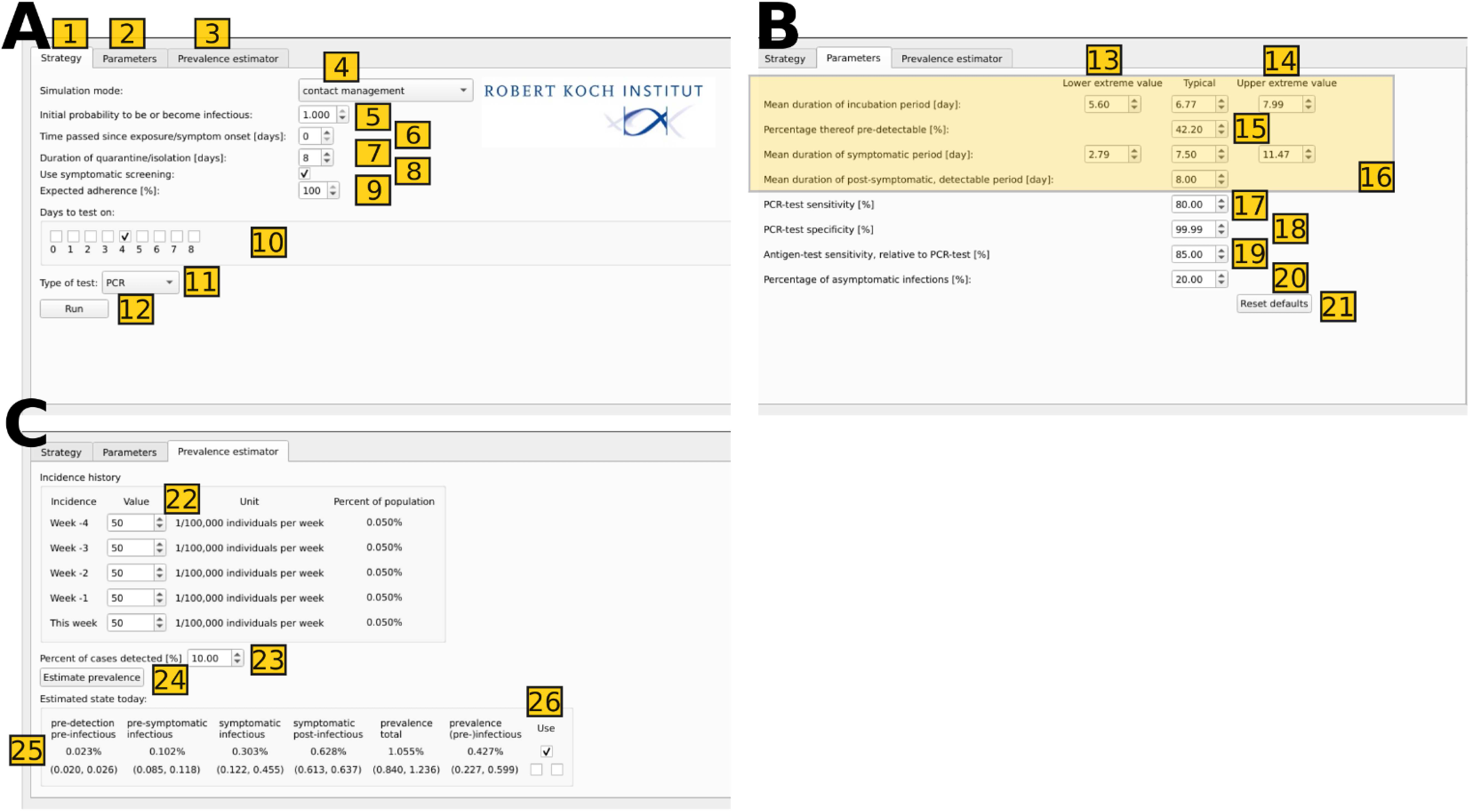
User input panels of the COVIDStrategyCalculator. **A.** Zoom-in on the NPI configuration window. **B**. Model parameter input tab. **C**. Prevalence estimator input tab. Details of the usage and meaning of the different input fields are provided in the text.’

### Configuring and simulating an NPI strategy

The COVIDStrategyCalculator has three *modes* of operation, corresponding to the three common scenarios in policy making: (i) quarantine for contact management, (ii) isolation of infected individuals and (iii) quarantine for incoming travelers. The modes differ in the initial states used for simulation. In the ‘contact management’ and the ‘isolation’ mode, the simulation starts from a point distribution, whereas in the ‘incoming travelers’ mode assumes a mix of ‘infection ages’.

(i) In the *contact management* mode, strategies aim to reduce the risk emanating from individuals who have been in recent contact with a confirmed case. In this scenario, the time of the putative infection is known and can be entered by the user (Fig. 3A 6.). In the underlying model (Fig. 1A), all states are set to zero, except the very first compartment of the model, which is set to the initial probability of infection provided in (Fig. 3A 5.). It is important to note here that the NPI efficacy estimates (‘relative risk’, ‘risk reduction’) are independent of the initial probability of infection, as they cancel out when computing these relative quantities (eq. (9)). The user selects a duration of quarantine (7.) and whether ‘symptom screening’ is performed (checkbox in 8.). ‘Symptom screening’ would imply that individuals who develop symptoms go into isolation (in line with WHO guidelines ^1,2,5^) and do not pose a risk anymore, whereas individuals that not develop symptoms are being released into society at the end of the quarantine and may continue to pose a risk. The percentage of *asymptomatic* infections who would be missed in ‘symptom screening’ can be provided by the user (outlined below). The expected level of adherence (= which percentage of individuals follows the proposed NPI) can be set in field (9.). The level of adherence *w* alters the risk calculation as follows:

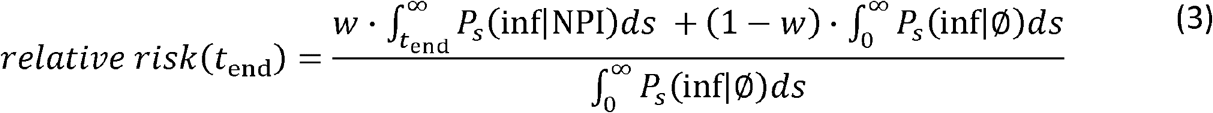

The user can also decide on whether diagnostic tests should be conducted during the quarantine (check-boxes in field 10.) and select whether PCR tests (default) or antigen-based rapid diagnostic testing (RDT) should be performed (field 11.). In case of a positive test, it is assumed that the individuals go into isolation and do not pose a risk anymore. Individuals that are false negative tested are assumed to stay quarantined until the end of the user-defined quarantine duration and are subsequently released into society. Therefore, the time-dependent *false omission rate* (= 1-sensitivity) of the test(s) is critical to determining the efficacy of testing during NPIs. Parameters related to the diagnostic test can be modified by the user as outlined below. The output of the configured NPI is shown immediately upon pressing ‘Run’.

(ii) In the *isolation* mode, the user can assess strategies for the duration of isolation after *symptom onset*. In this scenario, the time of symptom onset is known and can be entered by the user (Fig. 3A 7.). In the underlying model (Fig. 1A), all states are set to zero, except the very first ‘symptom compartment’, which is set to probability of infection provided in (Fig. 3A 5.). The user’s options are similar to the mode described above, with the exception that a ‘symptomatic’ screening is not possible.

(iii) When choosing the *incoming travelers* mode (field 4 in Fig. 3A), the user is taken to the prevalence estimation subroutine of the software, Figure 3C. The prevalence estimation subroutine can be used on its own, or to generate a population with mixed ‘infection age’ for the analysis of NPI strategies. The details will be outlined below. To use the mixed population, the user presses ‘estimate prevalence’ (Fig. 3C, field 24) to generate an initial probability distribution (depicted in Fig. 3C, field 25), which can be used (Fig. 3C, 26.) for simulation Figure 3A. After setting the initial probability distribution, the user can proceed as described above.

### Setting model parameters and hypothesis testing

In an associated article^6^, we detailed the parameter estimation procedure to derive default parameters that capture clinically observed temporal changes and variability in test sensitivities, incubation- and infectious periods, as well as times to symptom onset. However, a user may want to assess the efficacy of an NPI strategy that uses a novel diagnostic test, or is directed against a novel virus variant, which exhibits different viral kinetics. For this reason, COVIDStrategyCalculator allows users to adapt all parameters related to viral dynamics, diagnostics, as well as the proportion of asymptomatic cases. When switching to the parameters tab Figure 3A (2.), the user is taken to the window depicted in Fig. 3B.

The first 4 rows (shaded area in Fig. 3B) allow to alter the time course of infection, i.e. (i) the *average* incubation time (= time before symptom onset), (ii) the fraction of this incubation time, where the infected individual is not infectious and the virus is not detectable yet, (iii) the *average* duration of the symptomatic phase, where the individual is infectious and the virus detectable, (iv) as well as the *average* duration of the post-infectious phase, where the virus is detectable, but not infectious anymore. Changing these parameters will alter the efficacy of NPI strategies (e.g. how long the duration of a quarantine needs to be), the time-dependent sensitivity of the diagnostic assay (i.e. *when* tests would be most beneficial), as well as the efficacy of symptomatic screening during quarantine.

The next three lines (fields 17-19 in Fig. 3B) allow the user to alter the characteristics of the diagnostic tests performed, i.e. (v) maximum sensitivity of the PCR test (field 17.), as well as (vi) its specificity (field 18.). The (vii) sensitivity of antigen-based rapid diagnostic tests (RDT) is usually measured relative to PCR results ^13-16^ and can be set in field 19. of Fig. 3B. The temporal changes in test sensitivity for RDT are assumed to be comparable to those of PCR testing, but can be altered as described below. The RDT specificity is assumed to be equal to the PCR specificity, because, at least in low-prevalence / low pretest probability settings, a positive RDT result is typically confirmed by a PCR test ^17-20^. Generally, upon a SARS-CoV-2 diagnosis it is assumed in the COVIDStrategyCalculator that the individuals go into isolation and do not pose a risk anymore. Thus, altering diagnostic parameters (v-vii; fields 17-19) allows to change the efficacy of the tests to identify infected individuals, while the inputs (v-vii) shift the magnitude of the *false omission rate*. The temporal profile of the *false omission rate* can be altered by changing viral dynamics parameters (i-iv; fields in the shaded area in Fig. 3B). The resultant temporal test sensitivity can be inspected in Figure 2A (1.). Lastly, (viii) the proportion of asymptomatic cases can be changed in field 20 of Figure 3B. This parameter alters the effectiveness of ‘symptom screening’ in NPI strategies. Altering any of the parameters allows the user to perform hypothesis testing: For example, ‘How much worse would an NPI perform if the test sensitivity was lower?’, ‘… if the virus incubation time was shorter but the infectious phase longer?’ or ‘… if the proportion of asymptomatic cases was larger’, etc.

### Prevalence estimation and assessing NPI strategies in populations with mixed ‘infection age’

In COVIDStrategyCalculator, the user has the possibility to perform a prevalence estimation given a user provided incidence history of the past 5 weeks. The user can also use this utility to generate a population with mixed ‘infection age’ when computing NPI strategies in the ‘quarantine for incoming travelers’ mode.

To perform a prevalence estimation the user inputs the incidence history of the past 5 weeks in the region of interest (Fig. 3C, field 22). The incidence reports are typically reported as the number of cases per week and 100,000 inhabitants by national or supra-national health authorities, like the ECDC. In addition to the incidence reports, an estimate of the presumed proportion of cases that are actually reported (Fig. 3C, field 23.) can be provided. This number may vary widely between different regions and depends on national testing strategies. Upon pressing ‘estimate prevalence’, COVIDStrategyCalculator will estimate the prevalence based on the intra-host SARS-CoV-2 dynamics model (Fig. 1A), as detailed in an associated article^6^. The routine will calculate the probability distribution over the states of the model, as well as the ‘total prevalence’ and the prevalence of individuals that are currently *infectious*, or pre-*infectious*. Uncertainty ranges are calculated based on the extreme parameter values provided by the user (Fig. 3B, field 13-14.).

The estimated probability distribution can be used for assessing NPI strategies by checking ‘use’, which will take the user back to the NPI configuration window (Fig. 3A). While this feature of the software allows to estimate the prevalence in a setting of interest, it can also be used to set the initial distributions for the assessment of NPI strategies, e.g. by altering ‘incidence’ values to deduce a suitable initial distribution over the model states.

### Identifying efficient combined NPI + testing strategies

Another application of the COVIDStrategyCalculator is to identify NPI strategies that use testing to shorten quarantine or isolation periods. Essentially, this requires to find testing and quarantine strategies that are equivalent or non-inferior to established strategies. In order to identify those strategies, the user starts with assessing the efficacy (relative risk, risk reduction) of an established gold standard NPI as outlined in section “Configuring and simulating an NPI strategy” (e.g. 14 days quarantine as suggested by the WHO ^1^). Subsequently, the user could configure a combined strategy, where testing shortens the quarantine or isolation period, for example by placing a test at the end of the quarantine period. By repeating the procedure with different quarantine durations the user could find the shortest combined test + quarantine strategy which has a non-inferior efficacy (relative risk, risk reduction) to the gold standard NPI.

## Discussion

While vaccination programs are rolled out in most countries in early 2021, it is not yet clear when the critical level of vaccination will be achieved globally, or whether the SARS-CoV-2 virus will become endemic^21-23^. In both cases, non-pharmaceutical control strategies, including testing, isolation and quarantine will remain an integral part of controlling the further spread of SARS-CoV-2. We have developed an open-source software that allows decision makers to evaluate and deduce non-pharmaceutical SARS-CoV-2 mitigation strategies based on quarantine, testing and isolation. The software was designed to provide maximum flexibility to the user combined with intuitive operability. The web-version of the software runs in a platform-independent manner in the bowser (https://COVIDStrategyCalculator.github.io), while offline versions for all major operating systems are available for download from https://github.com/COVIDStrategyCalculator/COVIDStrategyCalculator. The underlying mathematical models and methods, which reproduce the spectrum of clinically observed infection dynamics from in-house and published studies, are presented in an associated paper^6^. As many public health experts may not be comfortable with implementing the underlying methods themselves, the main purpose of the software is to provide a tool that allows configuring custom NPI strategies and perform hypothesis testing for rational, evidence-based design of non-pharmaceutical control strategies.

An imminent question that may arise in the near future is whether NPI strategies need to be adapted in the context of increasing amounts of infections with novel, potentially more contagious variants^24-26^. For example, as of March 2021, the variant of concern (VOC) B.1.1.7 (also 202012/01 or 20B/501Y.V1 depending on nomenclature) is on the rise in the US, most European countries and has become the major circulating variant in the UK. Early reports ^27^ describe potentially distinct viral kinetics, based on 7 densely sampled individuals infected with B.1.1.7. While more data is required to ascertain potentially different viral kinetics in the light of the observed inter-individual variability, our software is already able to accommodate different viral kinetics, e.g. for hypothesis testing. For example, Kissler et al.^27^ suggest that the viral load rises and drops slower in B.1.1.7 compared to non-B.1.1.7, but achieves higher viral titers for prolonged durations pre- and post peak viral loads. In our software, these viral kinetic alterations can be implemented by increasing the ‘mean duration of incubation’, decreasing the ‘percentage thereof pre-detectable’ and increasing the ‘mean duration of the symptomatic period’ in the ‘parameters tab’ (compare section *Setting model parameters and hypothesis testing*). When we performed calculations with aforementioned parameter changes in COVIDStrategyCalculator, we generally observed that longer durations of quarantine would be needed to achieve risk reductions that are non-inferior to non-B.1.1.7. However, when combining quarantine with testing, the differences were less pronounced. In the future, it is planned to update the default parameter settings as soon as sufficient intro-host viral kinetic data for the novel variants is available.

Currently, many novel diagnostic options become available. Among these are antigen-based diagnostics for self-testing. The diagnostic performance of these tests is usually compared to PCR-testing of nasopharyngeal swabs^13-16^. In COVIDStrategyCalculator, it is straightforward to incorporate the relative sensitivity of these tests for hypothesis testing and for the design of NPI strategies that utilize self-testing, as outlined in section *Setting model parameters and hypothesis testing*.

Output parameters in the COVIDStrategyCalculator, such as the ‘relative risk’ or the ‘risk reduction’ are calculated with regards to the baseline risk of “no NPIs and unrestricted entry” (eq. (2)). Different baselines can also be computed based on the software outputs: One can compute the *relative risk* for intervention A and B respectively with regards to the default baseline ‘Ø’ in COVIDStrategyCalculator. Let us call them *RR*(*A*/Ø) and *RR*(*B*/Ø). The fraction 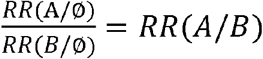 will give the *relative risk* of intervention A with regards to intervention B. Similarly, one may compute the *relative risk* of a fixed strategy in a variant of concern vs. a non-B.1.1.7 variant.

The COVIDStrategyCalculator allows to assess the efficacy of arbitrary NPI strategies with regards to the reduction in onwards transmission emanating from a (potentially) infected individual. Epidemiological modelling (e.g. SEIR) often addresses the question of disease spread in a population, as a consequence of different (hypothetical) interventions^28^, such as, for example, ‘how many individuals are expected to be infected, if a 96% effective vaccine was applied to 60% of individuals’, or ‘… if an NPIs strategy reduced transmission by 70% and was conducted by 50% of the individuals’. The efficacy parameters for these (hypothetical) interventions are often assumed. However, COVIDStrategyCalculator provides the required parameters of efficacy (‘relative risk’, ‘risk reduction’) that can be multiplied with default (‘no intervention’) parameters used in epidemiological models. The NPI efficacy estimates may also be used in conjunction with parameters of vaccine efficacy^29-31^ to evaluate the concomitant effects of both public health interventions.

In summary, we present COVIDStrategyCalculator, which is a free, platform independent, software that computes the efficacy of NPI strategies with regards to reducing SARS-CoV-2 transmission. The tool is based on a stochastic model that resembles the within-host viral dynamics and temporal test sensitivities. The software allows users to configure and evaluate arbitrary NPI strategies and can be used for hypothesis testing, e.g. with regards to designing NPIs to contain novel variants, or utilizing novel diagnostic tests into NPI strategies.

The web-version of the software can be used through https://COVIDStrategyCalculator.github.io and source-codes are freely available through https://github.com/COVIDStrategyCalculator/COVIDStrategyCalculator

## Data Availability

All data for reproducing the results is available as outlined in the manuscript.

https://github.com/CovidStrategyCalculator/CovidStrategyCalculator

## Experimental Procedures

### Resource Availability

Further information and requests for resources should be directed to and will be fulfilled by the Lead Contact, Max von Kleist.

### Materials Availability

This study did not generate new materials.

### Data and Code Availability

All datasets used during this study were previously published. The software is available at https://github.com/CovidStrategyCalculator/COVIDStrategyCalculator.

### Implementation

The software is as a standalone graphical user interface (GUI) that can either be run from a browser (https://COVIDStrategyCalculator.github.io) or offline for Windows, Mac and Linux. Source codes and pre-built executables are freely available from https://github.com/COVIDStrategyCalculator/COVIDStrategyCalculator. The tool itself is implemented in C++ using the Qt (Version 5.9.5) and Eigen (Version 3.3.7) library and is provided under the GNU LGPLv3 license. The web-version uses Qt for WebAssembly.

## Working group on SARS-CoV Diagnostics at RKI

Sandra Beermann, Sindy Böttcher, Brigitte Dorner, Ralf Dürrwald, Max von Kleist, Janine Kleymann-Hilmes, Stefan Kröger, Martin Mielke, Andreas Nitsche, Djin-Ye Oh, Janna Seifried, Sebastian Voigt, Thorsten Wolff

